# Combining vaccination with *w*Mel for dengue control in Brazil

**DOI:** 10.64898/2026.07.12.26357821

**Authors:** Emilie Finch, Angkana T. Huang, Clare McCormack, Anna Vicco, Julio Croda, Neil M. Ferguson, Ilaria Dorigatti, Henrik Salje

**Affiliations:** Department of Genetics, University of Cambridge, United Kingdom; Saw Swee Hock School of Public Health, National University of Singapore, Singapore; MRC Centre for Global Infectious Disease Analysis and the Abdul Latif Jameel Institute for Disease and Emergency Analytics, School of Public Health, Imperial College London, United Kingdom; School of Medicine, Federal University of Mato Grosso do Sul, Campo Grande, Brazil; Oswaldo Cruz Foundation, Campo Grande, Brazil; Department of Epidemiology of Microbial Diseases, Yale School of Public Health, New Haven, CT, USA

## Abstract

Dengue virus is a severe threat to global health. Novel vaccine and *Wolbachia* technologies offer a potential path to dengue control; however, it remains unclear how to use these approaches in tandem. Using metapopulation transmission models, we simulated future dengue burden in Brazil. We estimated that *w*Mel releases in 300 municipalities could avert 12-22% of cases over the next 10 years, while a national vaccination campaign could avert 8-10% of cases in the total population and 39-45% within the vaccinated cohort. Projecting forward 40 years, we found *w*Mel releases would drive up the mean age of cases beyond increases due to the ageing population and increase population susceptibility to dengue. Finally, *w*Mel releases can help mitigate the future burden in southern Brazil where climate change is rapidly increasing dengue virus risk. These findings highlight the need to consider the two technologies in parallel to optimise efforts to combat future dengue burden.

## Introduction

Despite half of the global population living in areas at risk of dengue, effective dengue control has remained challenging^1^. Traditional vector control methods have failed to limit dengue virus (DENV) infection risk while vaccine development has been complicated by immunological interactions between four DENV serotypes and increased severity associated with a secondary infection^2,3^. Spatial patterns of DENV risk are also shifting, as climate change expands the geographical range of *Aedes* mosquito vectors^4,5^. However, new vaccines and *Wolbachia*-based control methods now offer a potential path towards dengue control, with both interventions being rapidly scaled up across DENV endemic regions. Understanding how to optimally implement these new technologies is non-trivial as each intervention has different limitations, particularly regarding their interaction with the dynamic immune profile of populations and their logistical constraints, such as the availability of vaccine doses or the viability of *Wolbachia* release programs outside urban settings^6^. Thus far, no empirical evaluation of combined use in the same setting has been conducted.

Global DENV vaccination efforts are centered around Takeda’s Qdenga vaccine (TAK-003) and Butantan-DV, (Butantan Institute/NIH). Qdenga is a tetravalent, live attenuated vaccine licensed in 42 countries^7^. The safety and efficacy of Qdenga were evaluated in a Phase III trial, enrolling over 20,000 4-16 year olds in eight countries across Asia and Latin America^8^. After three years, overall efficacy was found to be around 62% against virologically confirmed dengue and 86% against hospitalised dengue^8^. More than 18 million Qdenga doses have been administered between 2022 and 2025, including during an unprecedented dengue outbreak in Brazil in 2024 that resulted in over 6 million cases, where the vaccine demonstrated real-world effectiveness in adolescents^7,9,10^. The second vaccine is a single-dose, tetravalent, live attenuated vaccine that was initially developed by the NIH, and has since been licensed to MSD, Butantan and others. The vaccine licensed to Butantan (Butantan-DV) was approved by Brazil’s regulatory agency, Anvisa, in 2025^8^. Phase III data for Butantan-DV showed an overall vaccine efficacy of 67.3% against virologically confirmed dengue with an average of 3.7 years of follow up^12,13^. MSD are currently undertaking Phase III trials of their version of the vaccine. Qdenga and Butantan-DV both demonstrated heterogeneous protection against disease by baseline serostatus and by serotype in Phase III trials, with TAK-003 having highest protection against DENV-2, and Butantan-DV highest protection against DENV-1. For Qdenga, no efficacy was observed in seronegative individuals against DENV-3, and efficacy against DENV-4 was unclear due to low incidence^14–16^. In the Butantan-DV trial, efficacy against DENV-3 and DENV-4 could not be evaluated as no cases were observed^13^. Both vaccines also showed higher efficacy in individuals with previous exposure to DENV. The key role of pre-existing immunity means that the effectiveness of vaccines will depend on historic DENV circulation patterns. Vaccine effectiveness will also evolve over time, as efficacy wanes and immunity shifts in the population due to public health interventions and changing (usually lowering) birth rates^16^.

In parallel, a mosquito-focused intervention has been developed, where *Aedes aegypti* mosquitoes are infected with strains of *Wolbachia* bacteria which facilitate their own introgression into local mosquito populations through reproductive manipulation, and reduce the ability of infected mosquitoes to transmit dengue and other arboviruses^17^. A cluster-randomised controlled trial of releases of *Aedes aegypti* mosquitoes infected with the *w*Mel *Wolbachia* strain found 77.1% protective efficacy against virologically confirmed dengue in release zones^18,19^. In most settings, *w*Mel-infected mosquitos displace local mosquito populations where introduced above a certain initial frequency^20^, with successful implementation in countries across Latin America, Asia and Oceania^21–25^. Within Brazil, large-scale releases in Niterói and Campo Grande successfully established *Wolbachia* within mosquito populations and resulted in a reduction in dengue incidence over subsequent years^21,26^. However, in Rio de Janeiro intermediate levels of introgression were seen following large-scale release efforts, and the drivers of successful introgression are not yet clear^27,28^. As large numbers of releases are required to successfully penetrate the local mosquito population, it may be financially and logistically difficult to achieve Wolbachia introgression in rural, low population density areas. The viability of achieving high levels of introgression outside urban settings, and methods to perform introgression across a broader, more spatially heterogeneous landscape are also unclear.

To understand the optimal use of these two interacting intervention strategies, we need to consider population immunity and demography alongside considerations of how each intervention could be deployed. In this study, we use mathematical modelling to quantify potential public health impact of vaccines and *Wolbachia* releases in Brazil, the country with highest reported global dengue burden, where both technologies are being implemented^10,21^. We combine subnational estimates of the force of infection with demographic projections and efficacy estimates within a stochastic metapopulation model framework to simulate future dengue burden in each federative unit of Brazil. We then evaluate the public health impact of different intervention scenarios by estimating potential cases, hospitalisations, deaths and disability adjusted life years (DALYs) averted. Finally, we consider long-term implications of *w*Mel release strategies on immunity and public health burden, including in scenarios of shifting DENV risk due to climate change.

## Results

### Potential impacts of vaccination and *w*Mel over the next decade

We projected the DENV burden across the 27 federative units of Brazil using a stochastic metapopulation transmission model incorporating demographic projections (Methods). We compared a scenario with no interventions with the following: a vaccination scenario with 60% coverage, assuming waning over 10-years; a *w*Mel release scenario targeting 300 municipalities; a combined vaccination and *w*Mel release scenario. For vaccination scenarios we assumed that the vaccine only protected against symptomatic disease, that efficacy varied by serostatus, and that the campaign involved a 3 year catch up campaign targeting 10-16 year olds followed by routine vaccination of 10 year olds. For *w*Mel scenarios we assumed that successful *w*Mel introgression reduces R_0_ by 50% with 70% introgression of *w*Mel infected mosquitoes into local mosquito populations (Methods)^30,31^. We initially selected municipalities for *w*Mel roll-out by targeting those with high population-weighted density and historical force-of-infection estimates (Figure 1a). Figure 2a shows the municipalities selected for *w*Mel releases; the majority were in the Northeast region (n = 257) with remaining municipalities in the North region (n = 38), Central-West region (n = 3) and Southeast (n = 2).

**Figure 1.**
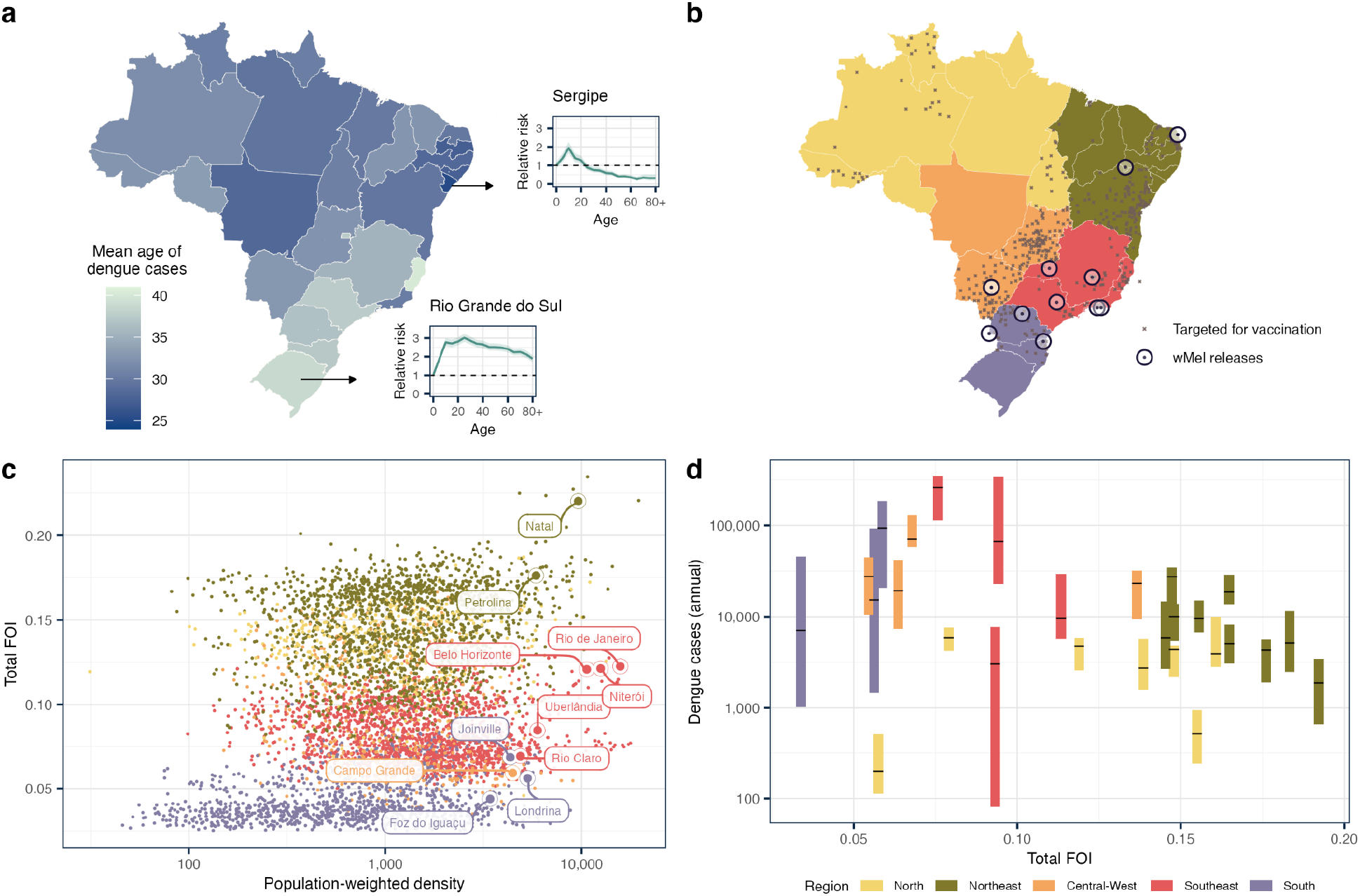
Dengue burden and interventions in Brazil. Panel a shows a map of the mean age of dengue cases in Brazil’s federative units according to SINAN data^53^. For two federative units (Sergipe and Rio Grande do Sul), the relative risk of being a dengue case by age are shown in subpanels (reference age = 1). Panel b shows municipalities that have been targeted for *w*Mel releases (black circles) and vaccination (grey points) with the underlying map coloured by region (North in yellow, Northeast in green, Central-West in orange, Southeast in pink and South in purple). Panel c is a scatter plot of municipalities in Brazil showing their population-weighted density (on the x-axis, with a log 10 scale) and total force of infection across all four serotypes (on the y axis). The points are coloured by region and municipalities which have already been targeted for *w*Mel releases are labelled. Panel d shows the distribution of annual dengue cases with bars showing the interquartile range and the black line showing the median. Bars are arranged by their estimated average force-of-infection^43,44^ and coloured by region.

**Figure 2.**
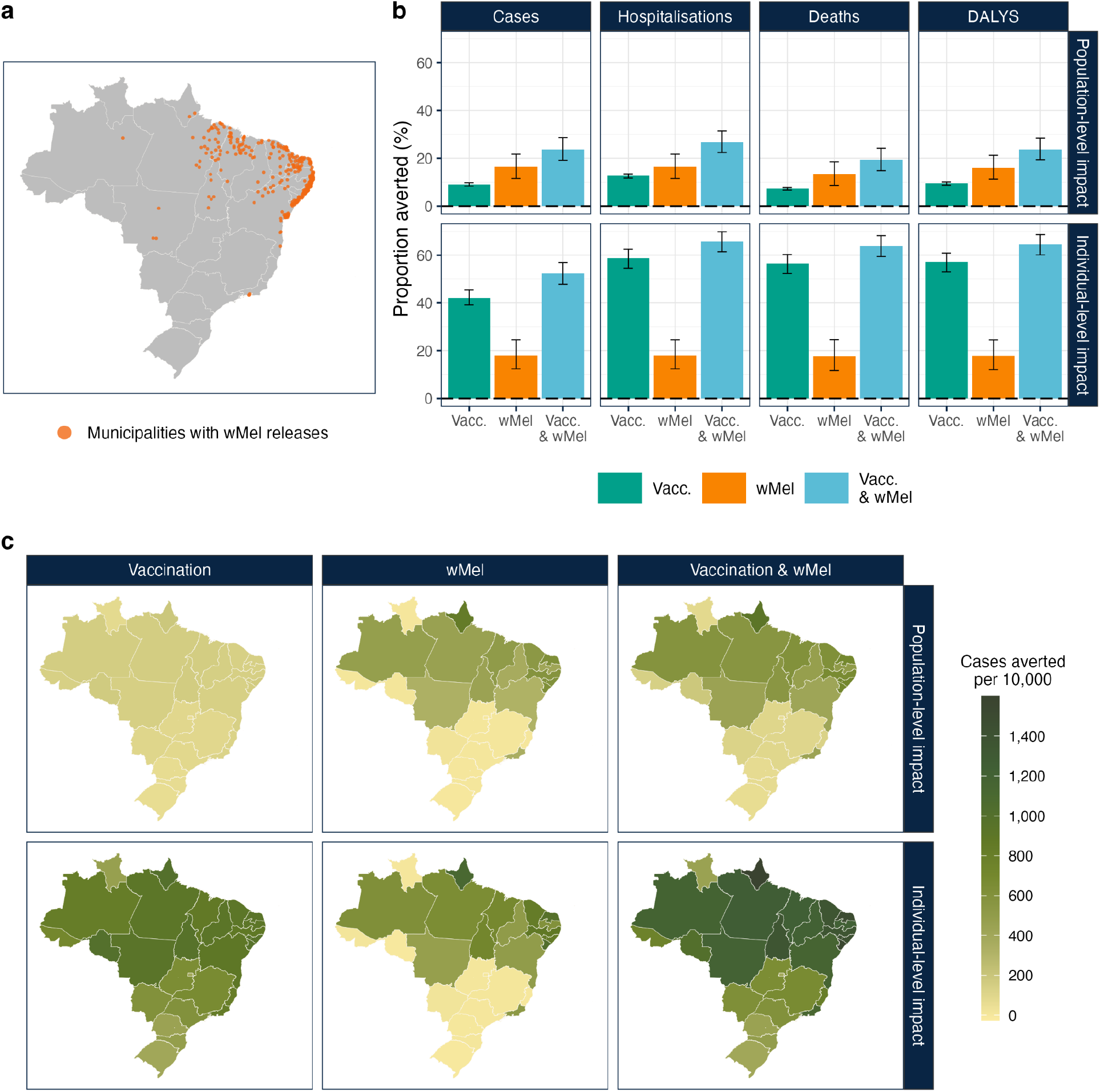
Public health impact of vaccination and *w*Mel releases over 10 years. Panel a shows the 300 municipalities targeted for *w*Mel releases in orange. Panel b shows the proportion of cases, hospitalisations, deaths and DALYS averted compared to a no intervention scenario for the following scenarios vaccination only (in green), *w*Mel only (in orange), vaccination & *w*Mel (in blue). The bars show the median proportion averted over 1000 stochastic simulations and the error bar shows the 95% uncertainty interval. Panel c shows the median cases averted per 10,000 population over 10 years by federative unit in Brazil over 1000 stochastic simulations. The top row shows population-level impact, that is the cases averted per 10,000 total population, while the bottom row shows individual-level impact, cases averted within vaccinated (or ‘would have been vaccinated’ cohort) per 10,000 vaccinated. From left to right, facets show a vaccination-only scenario, a *w*Mel-only scenario and a combined vaccination and *w*Mel scenario. Higher numbers of cases averted are shown in darker greens.

We then estimated the population impact of each intervention by calculating the public health burden averted in each scenario over 10 years compared to a no intervention baseline, estimated over 1000 stochastic model simulations. In a scenario with no interventions, we projected cumulative future burden of 26.8 million symptomatic cases over the next decade (95% uncertainty interval, UI: 25.9 - 27.8), alongside 2.4 million hospitalisations (95% UI: 2.3 - 2.5) and 58,000 deaths (95% UI: 56,000 - 61,000) (Supplementary Figure 7). We found that a *w*Mel campaign targeting 300 municipalities would avert around 16.4% of cumulative cases (95% UI: 11.5 - 21.7), while the vaccination campaign would avert 9.1% (95% UI: 8.4 - 9.8). Combining both interventions would increase this to 23.7% (95% UI: 19.2 - 28.7).

*w*Mel and vaccination offer two modalities of dengue protection. While the public health impact of *w*Mel would be expected to occur in release areas from the point of introgression onwards, the benefits of vaccination will accrue over time as more individuals are targeted by the vaccination campaign. Therefore, to enable a head-to-head comparison between the two interventions, we compared the burden averted per person year experienced under each intervention. Under our intervention assumptions, the *w*Mel campaign would target 46.8 million individuals, considering the population size in *w*Mel release municipalities in 2034, corresponding to 21% of Brazil’s population. Contrastingly, the vaccination campaign targets a median of 33.3 million individuals enrolled over the 10 year period, corresponding to around 15% of Brazil’s population. As individuals are enrolled sequentially into the vaccination campaign this translates to 256 million person years of protection while the *w*Mel campaign corresponds to 468 million person years of protection, while the combined scenario would include both (724 million person years). When adjusting for the person-years experienced under each intervention, we found that a *w*Mel campaign would avert 94 cases per 10,000 person years of intervention coverage (95% UI: 64 - 128), a vaccination campaign would avert 94 cases per 10,000 person years (95% UI: 87 - 103) and a combined campaign would avert 88 cases (95% UI: 69 - 109) per 10,000 person years (Supplementary Figure 8).

We also compared the impact of each intervention at the individual level, calculating the public health burden averted only in vaccinated individuals (or those that would have been vaccinated in counterfactual scenarios without vaccination). We find that the vaccination campaign would avert 42.2% of cumulative cases among this cohort (95% UI:39.2 - 45.4) while *w*Mel releases would avert 18.1% (95% UI: 12.5 - 24.5) and a combined strategy would avert 52.4% (95% UI: 47.8 - 57). Following our assumptions around *w*Mel and vaccination roll-out, the greatest impact of *w*Mel releases is concentrated in Northeast Brazil (in Amapá, Sergipe and Rio Grande do Norte). The impacts of vaccination are more evenly spread throughout the country but still higher in Northeast Brazil, where a high force of infection results in a lower average age of cases (Figure 2a).

Our results are reliant on several assumptions around dengue epidemiology in Brazil as well as the mechanism and roll-out of interventions. We therefore assessed how sensitive our findings are to assumptions around vaccination (efficacy, waning, target age and coverage), *w*Mel (introgression and number of municipalities targeted for roll-out) and dengue epidemiology (spatial coupling in transmission between regions and annual dengue introductions). We did this by varying each parameter in turn and calculating the percentage change in the estimated cases averted for each intervention scenario compared to our main results (Figure 3). We found that vaccine scenarios were most sensitive to assumptions around vaccine efficacy, target coverage and, to a lesser extent, vaccine waning and target age. *w*Mel scenarios were most sensitive to the assumed level of introgression into the local mosquito population and the number of municipalities targeted for releases. Finally all scenarios were sensitive to the assumed extent of spatial coupling in transmission between regions.

**Figure 3.**
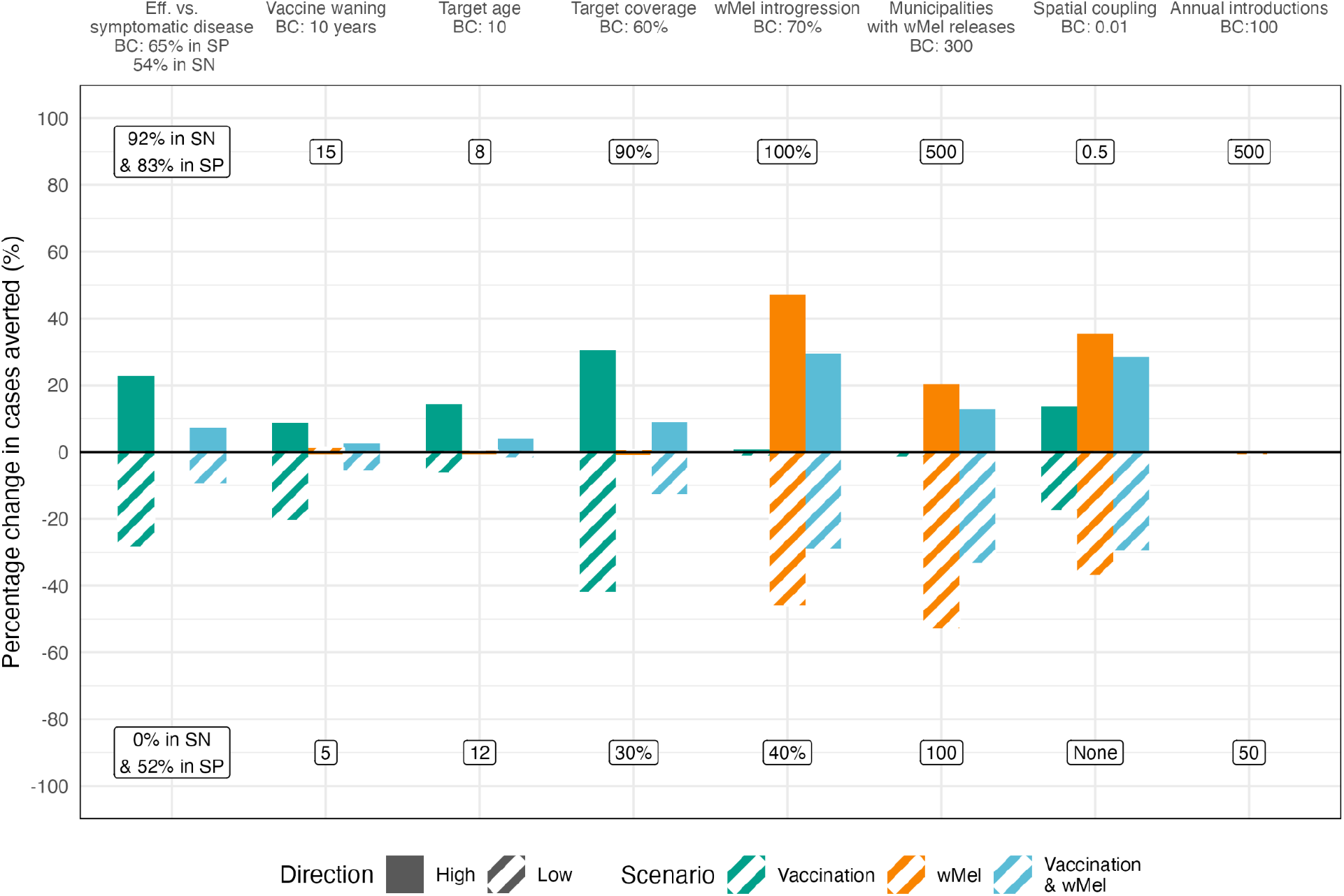
Sensitivity analysis of influence of model parameters on estimates of individual-level impact. Bars show the percentage change in cumulative cases averted at the population-level under different intervention scenarios when changing one parameter compared to the base case scenario. Full bars show the percentage change when the parameter is assumed to take a higher than in the base case, while striped bars show the percentage change when it is lower. Labels at the top of the facet panel show the ‘high’ value assumption while labels at the bottom of the facet show the ‘low’ value assumption. The percentage change for each scenario in the main analysis is shown including vaccination (in green), *w*Mel-only (in orange), and vaccination & *w*Mel (in light blue). Eff: efficacy; BC: base case; SN: seronegative; SP: seropositive.

### Long-term impacts of interventions on immunity

We then considered the potential longer-term impacts of interventions on population immunity and subsequent impacts on dengue transmission dynamics by generating projections of disease burden until 2065. In *w*Mel release areas, we found that the proportion of the population who are DENV-naive (or seronegative) increases in both *Wolbachia* scenarios (*w*Mel or Vaccination & *w*Mel) from around 7% in 2025 to 23% around 2050 (Figure 4a). At this point, outbreaks resume across *w*Mel release areas, and the proportion DENV-naive decreases, as enough population susceptibility has accumulated to drive Rt above 1 in some regions. We found that, in the absence of any interventions, changes in the underlying age structure of the population drive the mean age of cases up from around 16 to 25 by 2065 (Figure 4b). In scenarios with *Wolbachia* releases, we found the mean age of cases increases further still, to around 34 by 2065, as reductions in transmission intensity mean individuals escape infection for longer. We find a corresponding decrease in the cumulative deaths averted per 10,000 from 2060 onwards, as infections are shifted to older age groups with a higher probability of death given hospitalisation (Figure 4c).

**Figure 4.**
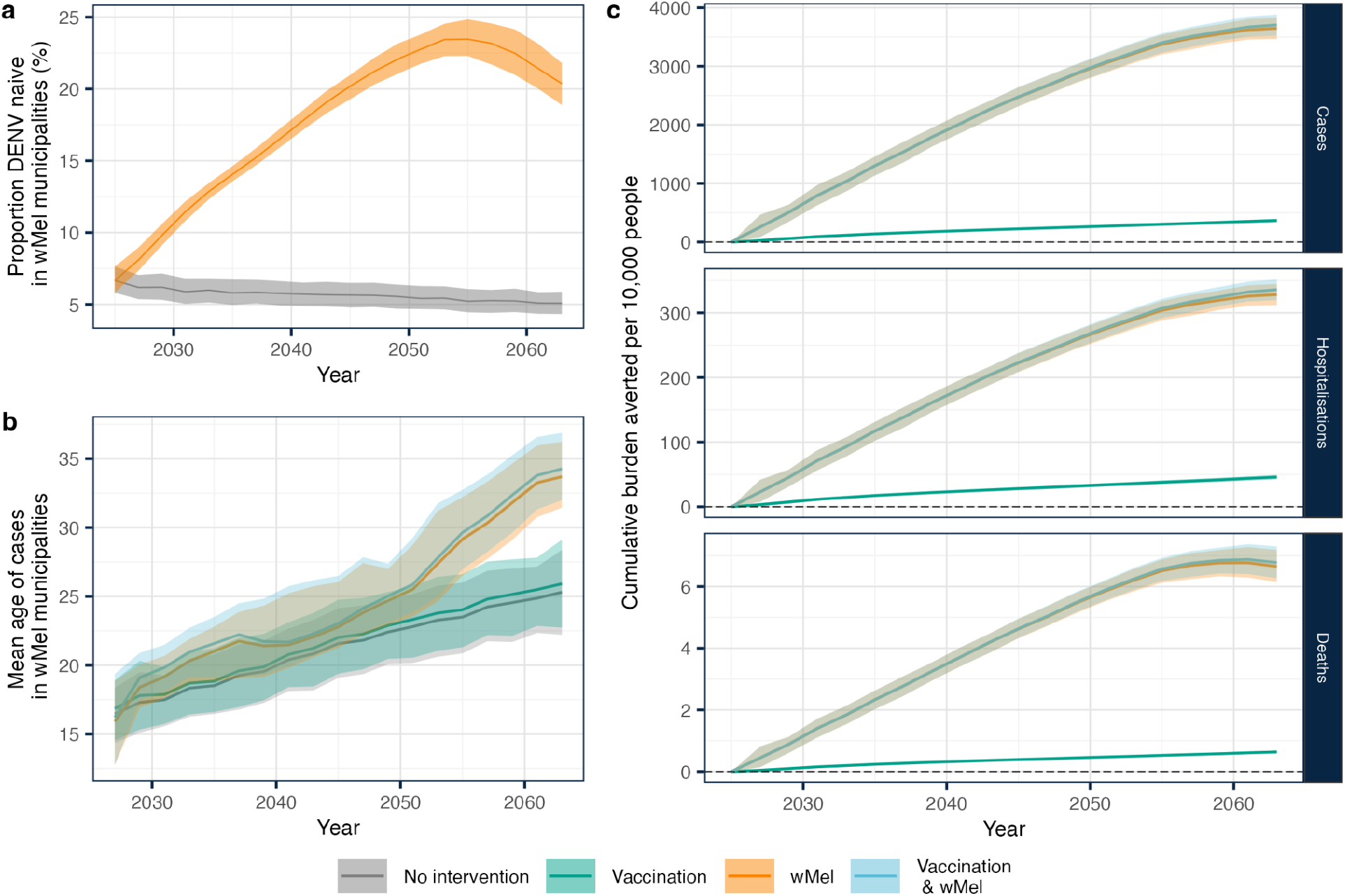
Long-term impacts of *w*Mel releases on immunity and public health burden in release areas. Panel a shows the proportion of individuals living in areas targeted for *w*Mel releases who are DENV naive (seronegative) in a no intervention and *w*Mel scenario. Note that, as we model vaccination as protecting against symptomatic disease and not infection, the proportion DENV-naive in the vaccination only scenario and vaccination and *w*Mel scenario would be identical to the no intervention scenario and *w*Mel only scenario, respectively. Panel b shows the mean age of cases in areas targeted for *w*Mel releases under each scenario. Panel d shows the cumulative cases, hospitalisation and deaths averted per 10,000 population in *w*Mel release areas.

### Evaluating alternative *w*Mel release strategies under expanding zones of transmission

Global heating is expanding the climate suitability for *Aedes* vectors and dengue transmission, shifting patterns of dengue transmission across Brazil and increasing outbreaks in regions that were historically protected, such as southern Brazil. *w*Mel releases are likely to have different implications in regions where dengue is newly expanding, where population susceptibility is high, than in historic hyperendemic regions. We evaluated potential impacts of future changes in R_0_ in southern Brazil on optimal *w*Mel release strategies, considering three different scenarios; no change in R_0_, a 20% increase in R_0_ over 10 years, and a 50% increase in R_0_ over 10 years. For each climate change scenario, we quantified the potential impact of alternative *w*Mel release strategies until 2065. The first strategy prioritises areas with high historical force of infection and high population-weighted density (as in previous analyses), while the second prioritises areas with high susceptibility and high population weighted density (Figure 5a). When considering the national burden averted per 10,000 population covered by *w*Mel releases in a scenario with no change in R_0_, we found that both strategies initially averted similar burden per-head. In the medium-term, a strategy targeting areas with high historical force of infection averted more cases than a strategy targeting highly susceptible regions. However, the reverse pattern is seen with deaths as individuals in susceptible regions are likely to be infected at older ages, with a higher associated risk of severe outcomes. Under scenarios of increasing R_0_ due to climate change, a strategy targeting areas of high susceptibility and population-weighted density is also impactful in the first 10-15 years as it helps to avert burden caused by large outbreaks seen as transmission increases in regions with low population immunity.

**Figure 5.**
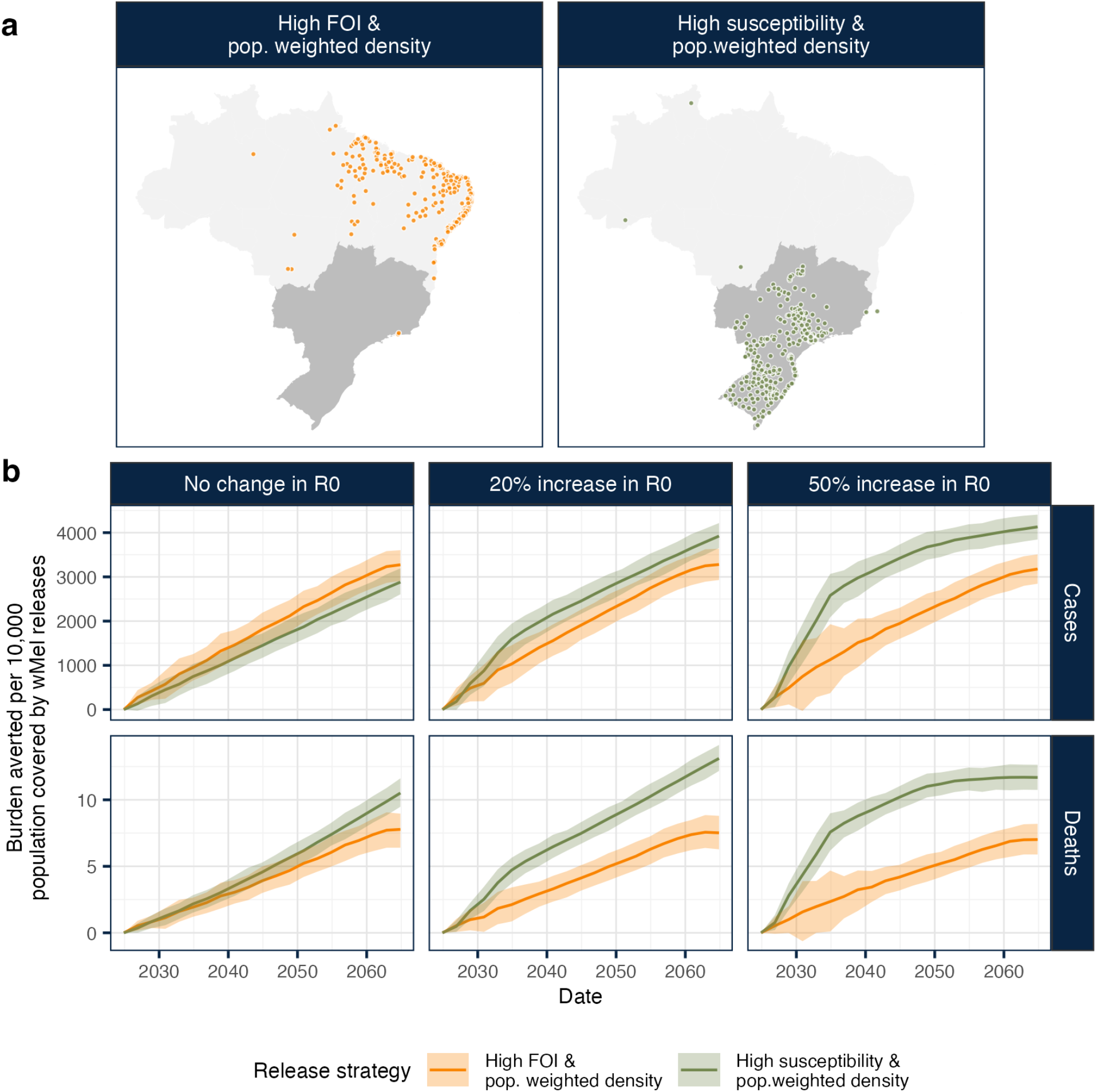
Comparison of *w*Mel release strategies under expanding zones of transmission. Panel a shows the top 300 selected municipalities for *w*Mel releases under two different rankings prioritising those with; 1) high force of infection (FOI) and population-weighted density, (in orange) 2) high susceptibility and high population-weighted density (in green). Panel b shows potential burden averted nationally for each *w*Mel release strategy considering scenarios with an increasing R_0_ in southern Brazil due to climate change (federative units in southern Brazil are highlighted in grey in Panel a). The columns show scenarios with; no change in R_0_, a 20% increase in R_0_ occurring over 10 years; a 50% increase in R_0_ occurring over 10 years. The rows show the cases and deaths averted from top to bottom.

## Discussion

We integrated estimates of dengue transmission intensity and demographic projections within a mathematical modelling framework to quantify the potential impacts of vaccination and *w*Mel releases on dengue burden in Brazil in the 21st century. We found that the differential impact of *w*Mel and vaccination over the next decade depended on whether we considered population-level or individual-level effects. At the population-level, we found a *w*Mel campaign targeting 300 municipalities could avert 12-22% of cases over the next 10 years, while a national vaccination campaign with 60% coverage could avert 8-10%. Combining both interventions increased the percentage of cases averted to 19-29% but resulted in a lower median number of cases averted per person-year compared with a *w*Mel only scenario. At the individual-level we found that the vaccination campaign averted 39-45% of cases amongst the cohort of vaccinated individuals.

We estimated that a national vaccination campaign and a *w*Mel campaign targeting 300 municipalities would avert a similar number of cases per person-years experienced. As such, our findings demonstrate the importance of understanding the relative costs and logistical constraints of scaling up *Wolbachia* releases and vaccination efforts to inform which intervention would be more appropriate or cost-effective for a given setting. For instance, as *Wolbachia* releases may be costly and logistically challenging to implement in rural areas, these settings could be prioritised for vaccination, which could be scaled-up more easily through school settings or health-care centres.

Our findings also highlight the complex effects of *w*Mel strategies on dengue transmission and burden in the longer-term. When projecting forward to 2065 we found decreased transmission intensity in *w*Mel release areas resulted in increased population susceptibility and drove up the mean age of cases, over and above that expected from the ongoing reduction in birth rates in Brazil^32^. As older individuals face a greater risk of severe disease, hospitalisation and death when infected, transmission-reducing interventions that shift disease burden into older ages could worsen the burden of severe outcomes^33–37^. While vaccine efficacy is still under evaluation in older adults, if efficacious, vaccination of older age groups could complement *w*Mel releases to reduce this risk.

By comparing alternative *w*Mel release strategies we found that the impact of different *w*Mel release strategies on dengue burden varied depended on the outcome considered (i.e. cases or deaths) and on future changes in R_0_ due to climate change. Climate change is shifting patterns of dengue transmission in Brazil leading to large outbreaks in areas with low levels of prior immunity, as seen in São Paulo in 2024^38^. We found that *w*Mel releases in susceptible regions could help mitigate these outbreaks. These findings highlight the importance of considering current seroprevalence and real-time estimates of underlying transmission intensity (R_0_) alongside population density when selecting target areas for *w*Mel releases^39^.

There is evidence that *w*Mel is sensitive to high temperatures, with heat waves potentially causing *w*Mel to ‘drop-out’ of the mosquito population^40,41^. Our estimates do not account for any temperature dependent effects on efficacy and further research is needed to characterise the relationship between temperature fluctuations, *Wolbachia* and dengue transmission to improve estimates of potential impact under different climate change scenarios. There is evidence that both vaccination and *w*Mel interventions show differing efficacy by serotype. TAK-003 has shown greatest efficacy against DENV-2 in both seropositive and seronegative individuals, with no evidence for efficacy against DENV-3 and DENV-4 in seronegative individuals^14,15^. The ability of *w*Mel to block DENV transmission is also thought to vary by serotype, with greatest blocking for DENV-2 and DENV-4 and weaker blocking for DENV-3^42^. We did not incorporate serotype-specific dynamics into our modelling framework; however, we assessed the sensitivity of our vaccination impact results to our assumptions around circulating serotypes by testing scenarios with vaccine efficacy estimates for DENV-2 and DENV-3 respectively. High levels of transmission of, for instance, DENV-3 strains would result in a lower burden averted by vaccination and potentially *w*Mel. Uneven inhibition of DENV strains by *Wolbachia* releases could result in selection for strains that are minimally inhibited or leave open the potential for outbreaks if a minimally inhibited strain is introduced^42^. Genomic viral surveillance in intervention areas will be crucial to understand the impacts of *Wolbachia* on DENV strain diversity and improve assessments of public health impact. Finally, we do not consider the impacts of heterogeneity in *w*Mel introgression, as seen in complex urban environments such as Rio de Janeiro^28^. This could reduce potential impacts of *w*Mel on public health burden or, equally, by allowing for lower levels of ongoing transmission, slow the accumulation of population susceptibility and shifting of age burden to older ages. Finally, we ignore the role of *Aedes* albopictus, which is not impacted by *w*Mel interventions but is a viable DENV vector and could have a role in maintaining future DENV transmission in Brazil.

Despite these limitations, we provide estimates of potential impacts of expanded vaccination and *w*Mel campaigns on dengue burden in Brazil over the next decade and illustrate potential longer-term impacts of *Wolbachia* interventions on transmission dynamics. We present a generalisable modelling framework that could be adapted for use in other dengue-endemic settings and show how modelling can be used to incorporate current uncertainties around the mechanism and effectiveness of interventions to quantify differential impacts of specific intervention scenarios. This framework can be used to support ongoing co-delivery of vaccination and *w*Mel in endemic settings, such as in Dourados, Brazil, which has begun Qdenga vaccination and plans to begin *w*Mel releases, and makes a strong case for *w*Mel releases in areas where DENV is newly expanding, such as São Paulo. As climate change expands the geographical range of DENV transmission and populations age globally, careful integration of *w*Mel releases and vaccination campaigns offer the potential to transform future dengue burden.

## Methods

### Data

We used published estimates of the force of infection (FOI) at the municipality (admin 2) and federative unit (admin 1) level ^43,44^. From these, we calibrated corresponding transmission coefficient (*β*) values which were used as a model input (Supplementary Figure 2 and 3).

We constructed a dataset of single year age-stratified, municipality-level demography data by combining population estimates and projections from the Brazilian Institute of Geography and Statistics (IBGE). To do this, we used population size estimates by single year age group and municipality available from 2000-2024^45^. We then extrapolated municipality level data using IBGE population projections available from 2025-2070, stratified by single year age group and federative unit, and using age-stratified national level UN projections from 2070-2100^45,46^. We aggregated population data by age group (using single year age groups from 0:80+), municipality and year.

We obtained dengue case data from Brazil’s surveillance system (SINAN) in 2023 to validate our baseline model behaviour (Supplementary Figures 4 and 5)^47^.

### Selection of *w*Mel release sites

In this analysis, we selected 300 municipalities for *w*Mel releases to understand potential impacts of operationally realistic releases (i.e. not country-wide) on dengue burden. We selected municipalities based on two metrics: population-weighted density and force of infection estimates. Population-weighted density measures the population density experienced by an average person living within a geographical region and is computed by taking a weighted average of the population density in each 1×1 km^2^ grid square of a municipality, weighted by population size.

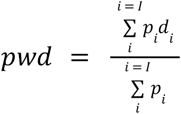

We normalised each municipality’s population-weighted density and estimated average force of infection using min-max scaling and then calculated a joint metric for each municipality using the product of their normalised population-weighted density and force of infection, selecting the top 300 for *w*Mel releases.

### Transmission model

#### Model equations

To quantify the potential impacts of vaccination and *w*Mel releases on dengue burden, we developed a stochastic metapopulation transmission model using *odin*.*dust*^48^. The model has an S-I-C-R framework, where the population is divided into susceptible, infected, cross-protected and recovered compartments. The model is stratified by age (using single year age groups from 0:80+) and federative unit. Individuals can be infected four times, once with each serotype, after which they experience lifelong homotypic immunity and cross-protection from infection by other serotypes with an average duration of one year.

Within our model, each federative unit is split into two areas - those with *Wolbachia* releases and those without. We then split the total population of the federative unit between these two areas, to reflect the population size of municipalities selected for Wolbachia releases. We incorporate spatial coupling in transmission between the 54 regions in our model (two regions for each of the 26 states and one federal district) using a spatial kernel. This is parameterised according to a gravity model, whereby *d* _(*m,n*)_ quantifies the spatial interaction from state *m* to state *n* such that:

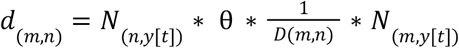

Here, *D*_(*m,n*)_ is the distance in km between state centroids, *N* is the state population size for year *y* and *θ* is the spatial coupling parameter. We assume that transmission scales linearly with the population at the source and the destination. This spatial kernel is then used to calculate a gravity-weighted force of infection exerted by other states on state *n* where:

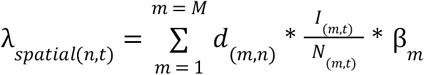

The total force of infection experienced by state *n* is then given by:

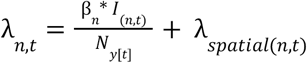

We used a discrete time stochastic modelling framework, with the deterministic dynamics described by the equations below. Here *δ*_*x,y*_ is the Kronecker delta function which is equal to 1 when *x* = *y* and to 0 otherwise.

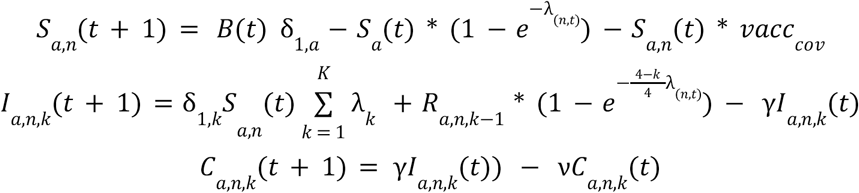

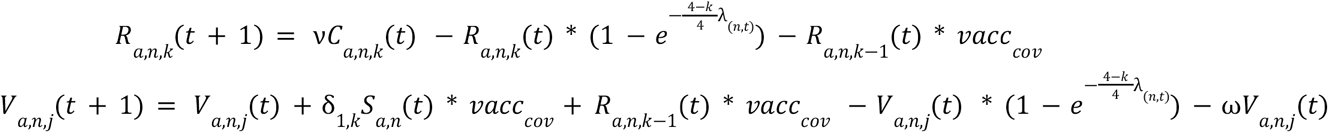

Here, all compartments are stratified by age *a*, region *n*. Compartments I (infected), C (cross-protected) and R (recovered) are also stratified by number of previous infections *k* which can take values 1 - 4. V (vaccinated) is also stratified by infection history, *j*, which can take levels 1 to 5, where j = 1 indicates individuals vaccinated while seronegative and j = 2 to 5 indicates individuals vaccinated with 1 to 4 previous infections prior to vaccination, respectively. *B*(*t*) represents the number of births, *vacc*_*cov*_ represents the proportion vaccinated, represents the probability of waning, *γ* represents the loss of infection/infectiousness and *v* represents the loss of cross-protection. Parameter values are shown in Supplementary Table 1.

Cases, deaths and hospitalisations were modelled as observation processes. We assume the probability of becoming a symptomatic case once infected varies by infection number with the second infection resulting in the highest probability of symptomatic disease. We assume that the probability of hospitalisation given that an individual has developed symptoms is 0.09 (regardless of infection number), in line with previous studies^49^. We assume an age-specific probability of death based on analysis of Brazil’s hospitalisation data (Supplementary Figure 1)^50^.

Ageing is modelled as a cohort process, with individuals moving up an age class once per year. At this point, the age class is scaled to match region and age-stratified demographic data (i.e. all the compartments for an age class are scaled upwards or downwards to align the total population size of the age class with the demographic data for that year). The total number of births for the year (the population size at *a* = 0) are spread out to occur evenly throughout the year and we do not incorporate any maternal immunity.

#### Vaccination

We modelled vaccination through the inclusion of a vaccine compartment (V). Vaccination occurred once a year with individuals in the eligible age cohorts moving to the V compartment with a probability corresponding to the assumed vaccine coverage.

We used vaccine efficacy estimates against symptomatic dengue (virologically confirmed dengue) and hospitalisation (hospitalised virologically confirmed dengue) from the phase III trial data of TAK-003 after 3-years^14^. These were 62% (95% CI: 56.6 - 66.7) and 54.3% (95% CI: 41.9 - 64.1) for seropositive and seronegative individuals against symptomatic disease, respectively, and 86% (78.4 - 91.0) and 77.1% (58.6 - 87.3%) for seropositive and seronegative individuals against hospitalisation, respectively. We assumed vaccine efficacy against death was the same as vaccine efficacy against hospitalisation. To incorporate uncertainty around vaccine efficacy, we fit a beta distribution to the mean and 95% confidence intervals of efficacy estimates and then drew 1000 samples for each vaccine efficacy parameter used, one for each stochastic simulation (Supplementary Figure 6). Density plots of the vaccine efficacies used are shown in Supplementary Figure 6. As estimated vaccine efficacy of TAK-003 varies by serotype, which we do not incorporate in our main analysis, we conducted sensitivity analysis around this using estimated efficacies against DENV-2 (against which protection is strongest) and DENV-3 (against which it is weakest) to understand how different serotype circulation could affect our impact estimates.

#### wMel releases

We modelled the impact of *w*Mel releases as a reduction on transmission (that is, on *R*_0_). In our main analysis, we selected 300 municipalities for *Wolbachia* releases and divided the population at the federative unit level into *Wolbachia* and non-*Wolbachia* areas as it would have been computationally infeasible to run the metapopulation model at the municipality level. We assumed that *w*Mel reduces *R*_0_ by 50%^30,31^. We then assume 70% introgression of *w*Mel into mosquito populations and modelled this by assigning 30% of the population within each federative unit residing in *Wolbachia* areas to non-*Wolbachia* areas. We conducted a sensitivity analysis around our assumptions around the number of municipalities targeted for releases and introgression levels in a sensitivity analysis.

### Counterfactual comparisons

We equilibrated the model over 150 years, assuming static demography before 2000, and ran 1000 stochastic equilibration runs to use as initial states for onward simulation. We then simulated 1000 stochastic simulation for the following scenarios:

1. No intervention scenario
2. *w*Mel scenario with releases in 300 municipalities, assuming 70% introgression and 50% reduction in *R*_0_
3. Vaccination scenario with 60% coverage. We model a nation-wide 3-year catch up campaign targeting 10-16 year olds followed by routine vaccination of 10 year olds. We assume vaccination is disease-blocking with vaccine efficacies against disease, hospitalisation and death shown in Supplementary Figure 6
4. Combined *w*Mel and vaccination scenario

Within non-vaccine scenarios, individuals were still moved to the vaccine compartment but with vaccine efficacies of zero, enabling us to compare the impacts of each intervention at the population- and individual-level, following Cracknell Daniels et al^16^. The population-level impact compares the number of cases, hospitalisation, deaths and DALYs averted within the entire population (including both vaccinated and non-vaccinated individuals). Individual-level impact was calculated as the burden averted within the vaccinated cohort (or those that ‘would have been vaccinated’ in non-vaccine scenarios). We also considered the public health impact over different time periods, considering impact over a 10-year and 40-year time horizon.

To estimate the burden averted by each intervention, we calculated the number of cases, hospitalisations, deaths and DALYs averted for each run between the intervention scenario and a baseline scenario with no interventions at a national and regional (federative unit) level. We also calculated the proportion of burden averted in each run. We then calculated the median number and percentage of cases, hospitalisation, deaths and DALYs averted, alongside and 95% uncertainty quantiles. DALYs were calculated using IGBE life tables from 2023^51^ with weights of 0.051 for symptomatic dengue and 0.133 for hospitalisation due to dengue^52^. The uncertainty interval (UI) associated with these estimates incorporates stochastic uncertainty, parametric uncertainty in the vaccine efficacy parameters and uncertainty in the initial conditions.

## Data availability

All code and data used for this analysis are available at: https://github.com/EmilieFinch/brazil-denv-vacc-wmel

