## Supplementary Materials for "Combining vaccination with *w*Mel for dengue control in Brazil"

| Parameter | Description | Value | Reference |
|---|---|---|---|
| β | Transmission coefficient | Calibrated to estimates of average annual force of infection by federative unit (see Supplementary Figure 2) | |
| $\gamma$ | Rate of recovery from infection (1/duration of infection) | 0.2 | [1–3] |
| $\nu$ | Duration of cross-protection | 1 year | [4] |
| p(symptoms\| infection) | Probability of apparent infection given infection number | Primary infection = 0.214<br>Secondary infection = 0.583<br>Tertiary infection = 0.288<br>Quaternary infection = 0.288 | [5] |
| p(hospitalisation \| symptoms) | Probability of hospitalisation given symptomatic disease | 0.09 | [6,7] |
| p(death \| hospitalisation) | Probability of death given hospitalisation | Age-specific (see Supplementary Figure 1) | [8] |
| θ | Spatial coupling parameter | 0.01 | Exploratory analyses |
| ω | Rate of vaccine waning | 1/(365*10) | Reflecting an average duration of protection of 10 years |

Supplementary Table 1: Parameter values

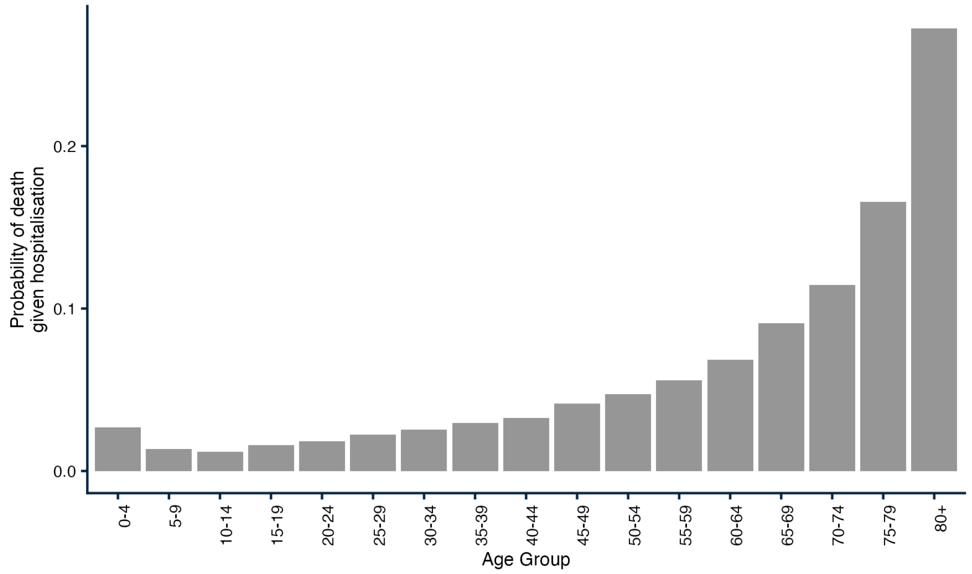


Supplementary Figure 1: Probability of death given hospitalisation used in model simulations, derived from Brazil's SUS Hospital Information System data[8]

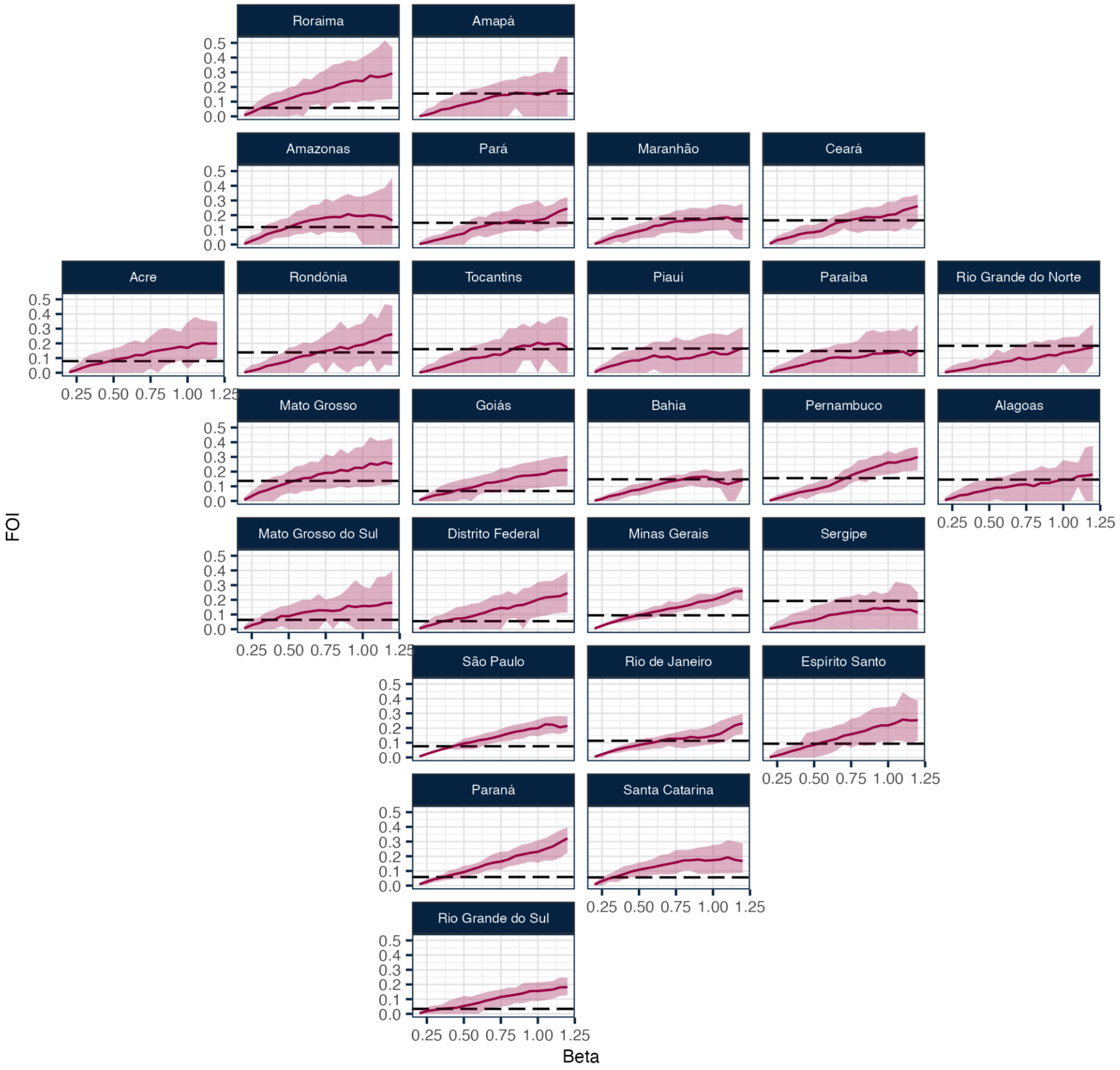


Supplementary Figure 2: Relationship between transmission coefficient (β) and average annual force of infection. Pink lines represent the average annual force of infection by federative unit after an equilibration period of 150 years for a given underlying β, averaged over 100 stochastic simulations. The pink shaded area shows the 95% uncertainty interval. The dashed line shows the published estimates of the average annual force of infection used to choose input β values for onward simulation[9].

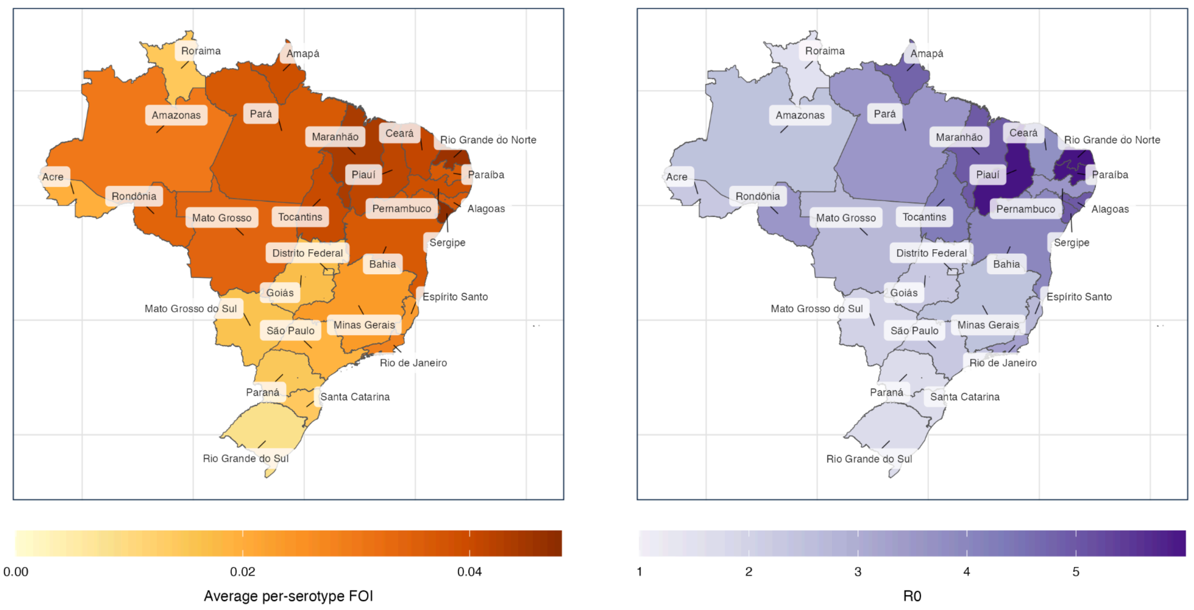


Supplementary Figure 3: Force of infection and corresponding $R_0$ in Brazil at the federative unit level.

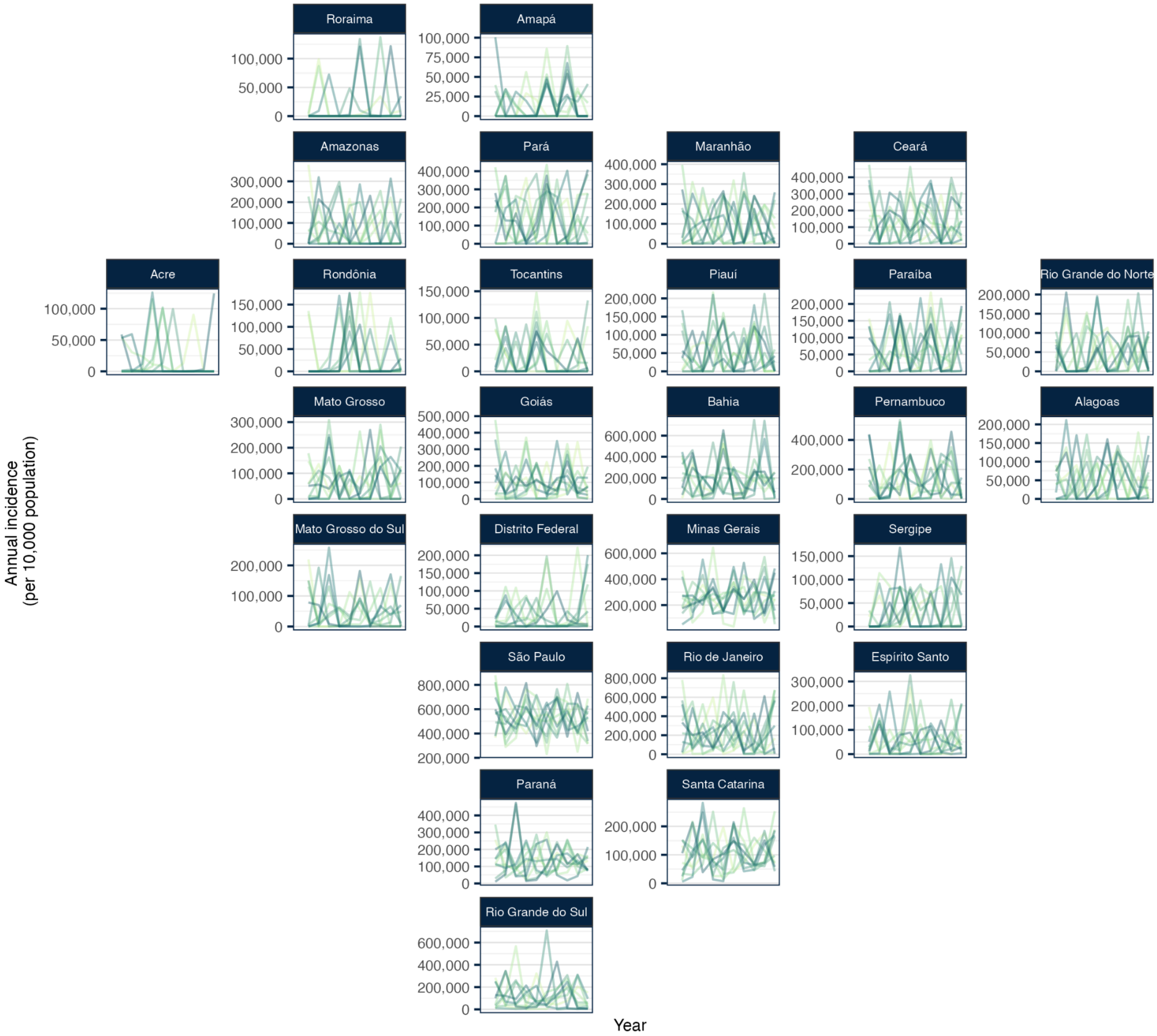


Supplementary Figure 4: Simulated dengue incidence over 10 years with no interventions. Green lines represent annual dengue incidence per 10,000 population for 10 randomly sampled runs from the no intervention scenario simulations in the main analysis.

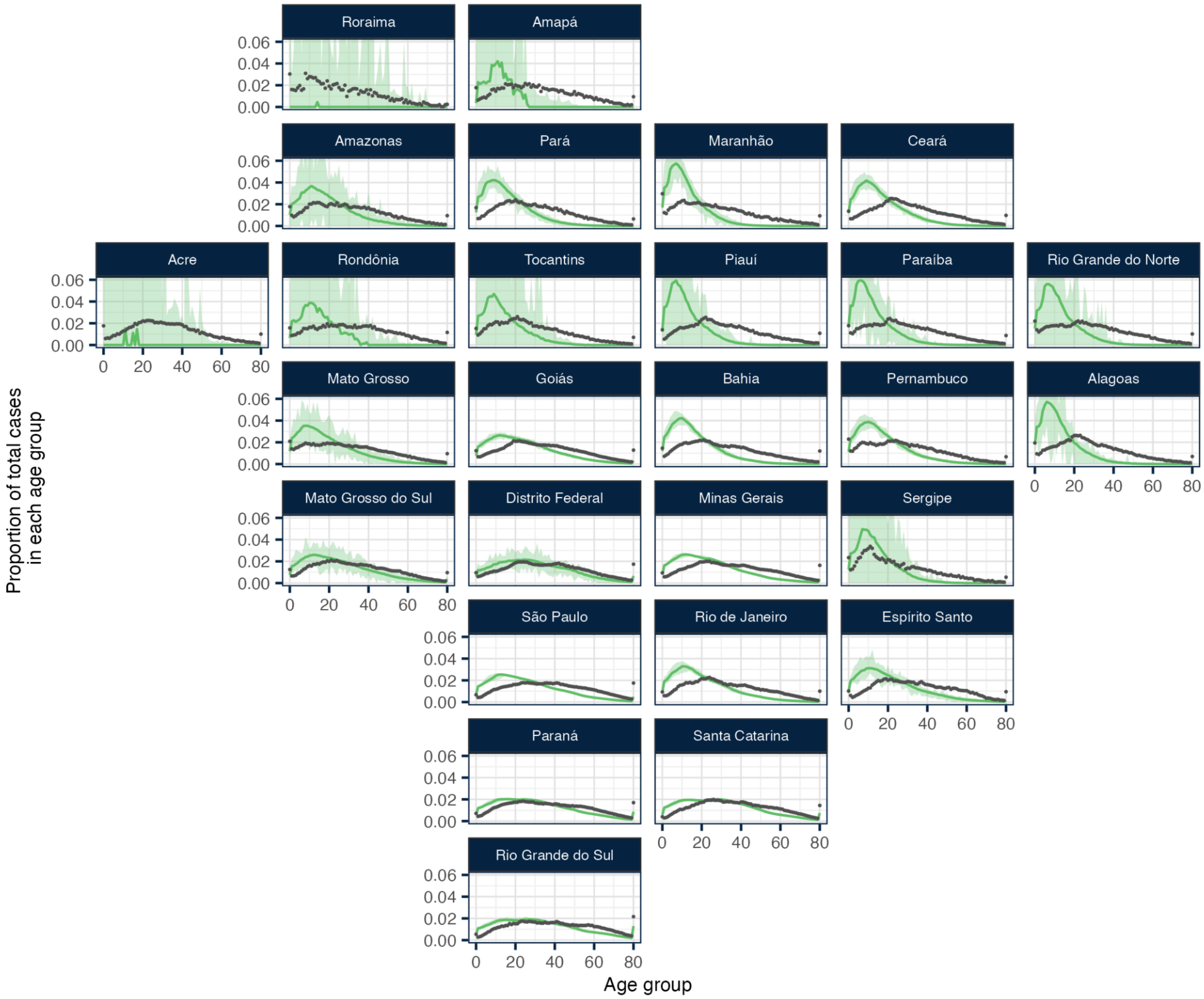


Supplementary Figure 5: Comparing the proportion of total cases in each age group from model runs with no interventions and SINAN surveillance data from 2023. Green lines represent the median proportion of total cases in each age group across 10 randomly sampled runs from the no intervention scenario in the main analysis, with the 95% uncertainty interval shown with a light green ribbon. Black points show the proportion of total suspected dengue cases in each age group from Brazil's surveillance data in 2023[10].

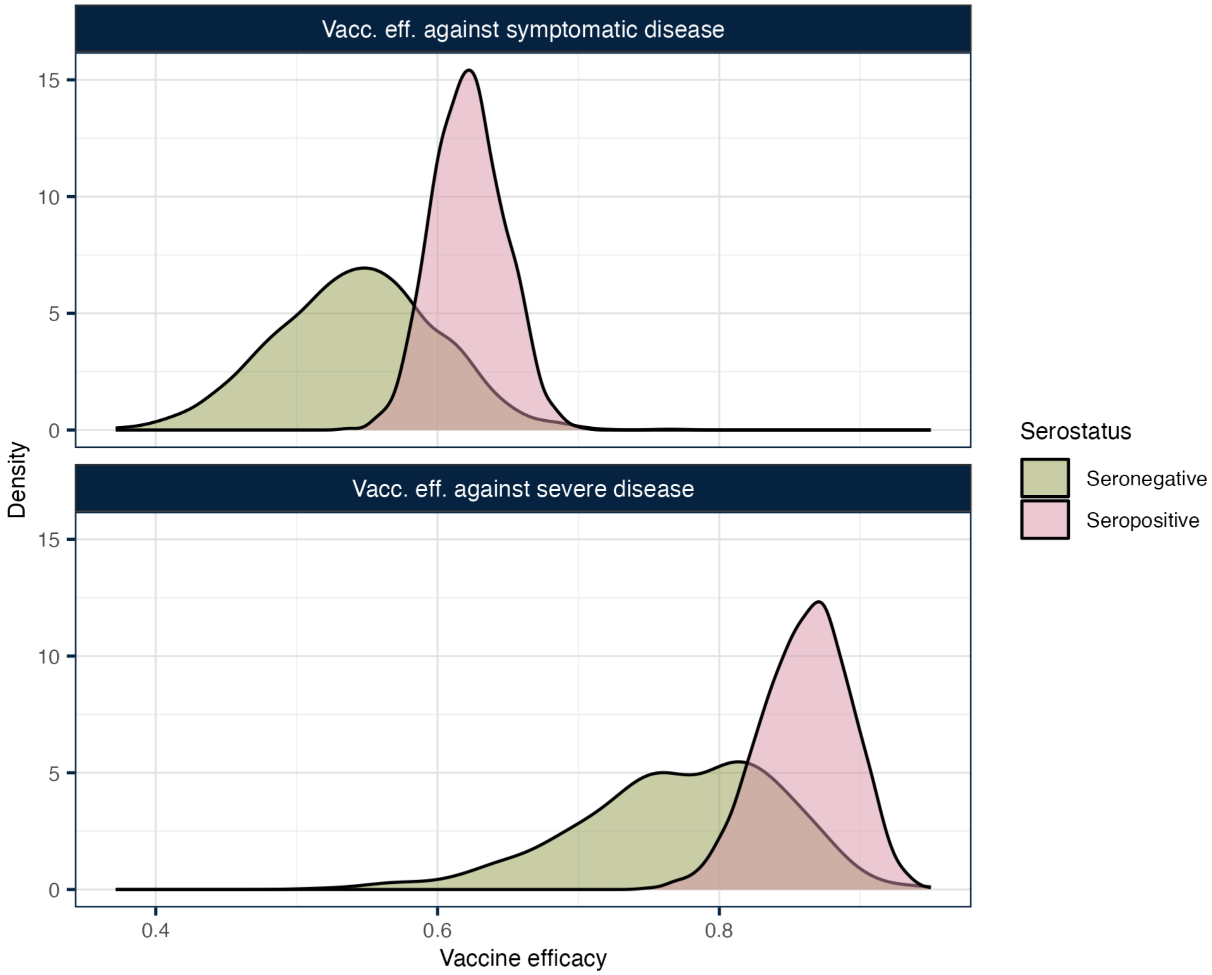


Supplementary Figure 6: Density plots of the vaccine efficacy samples used in model simulations. The figure shows the distribution of 1000 samples drawn from a beta distribution fit to the mean and 95% confidence intervals reported from the 3-year end point  phase III trial data from TAK-003[11]. Vaccine efficacy against death was assumed to be the same as vaccine efficacy against severe disease.

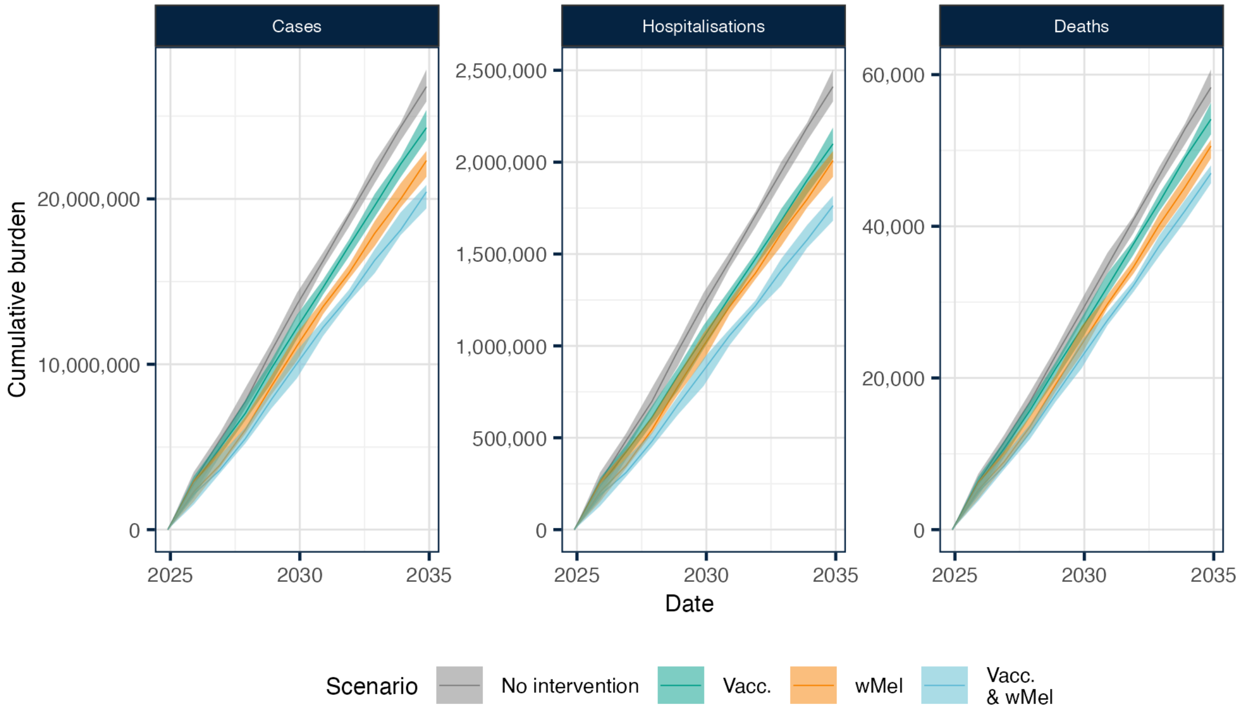


Supplementary Figure 7: Cumulative burden over 10 years. Panels show cumulative cases, hospitalisations and deaths during the simulation period for each scenario including: no intervention (in grey), vaccination (in green), wMel-only (in orange), and vaccination & wMel (in light blue).

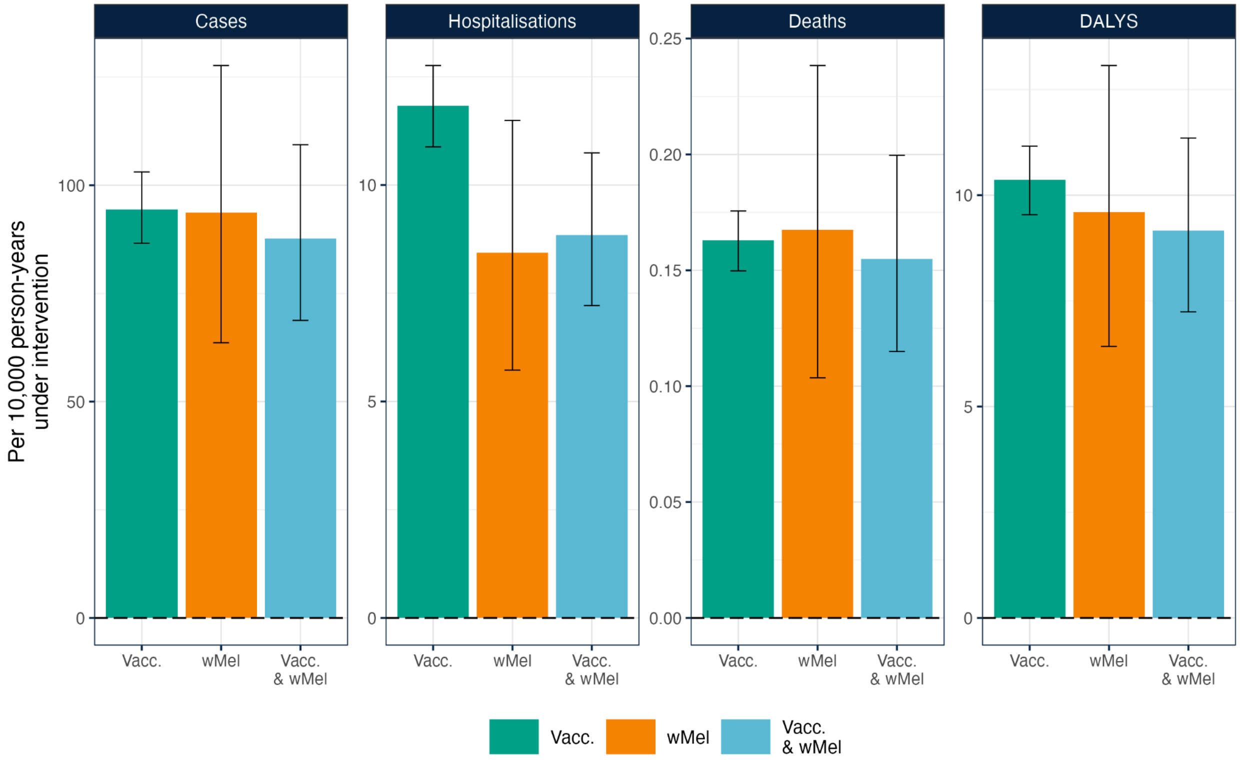


Supplementary Figure 8: Burden averted over 10 years per 10,000 person-years under intervention.
Figure shows the cumulative burden averted per 10,000 person-years experiencing each intervention for cases, hospitalisations, deaths and DALYS comparing: vaccination only (in green), *w*Mel-only (in orange), vaccination & wMel (in blue). Note here that we estimate that a *w*Mel campaign would cover 468 million person-years while a vaccination campaign would target 259 million person-years individuals, and a combined campaign would target the sum of both of these. The bars show the median burden averted over 1000 stochastic simulations and the error bar shows the 95% uncertainty interval.

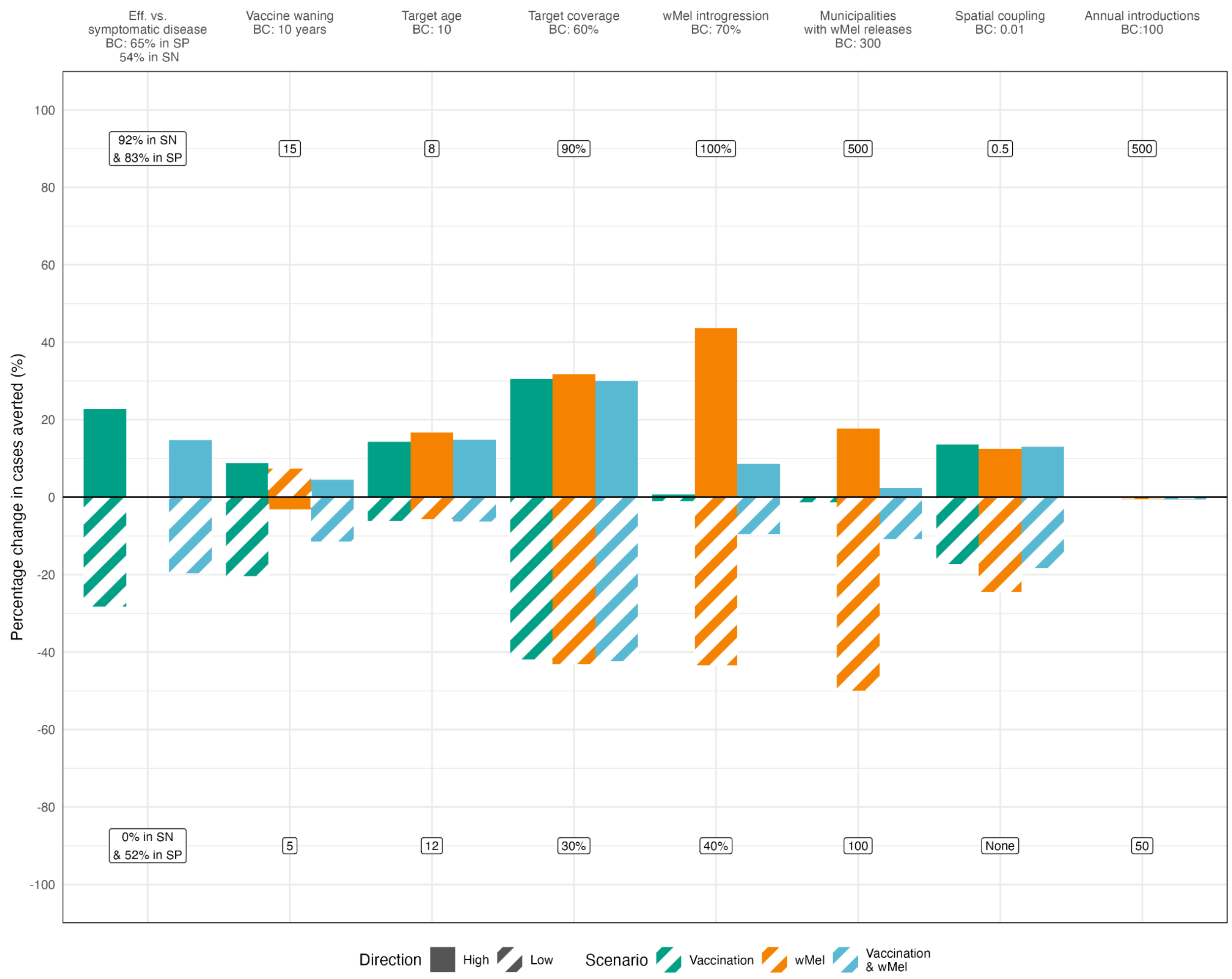


Supplementary Figure 9: Sensitivity analysis of influence of model parameters on estimates of individual-level impact

Bars show the percentage change in cumulative cases averted at the individual-level under different intervention scenarios when changing one parameter compared to the base case scenario. Full bars show the percentage change when the parameter is assumed to be higher than in the base case, while striped bars show the percentage change when it is lower. Labels at the top of the facet panel show the 'high' value assumption while labels at the bottom of the facet show the 'low' value assumption. The percentage change for each scenario in the main analysis is shown including vaccination (in green), *w*Mel-only (in orange), and vaccination & *w*Mel (in light blue). BC: base case; SN: seronegative; SP; seropositive.

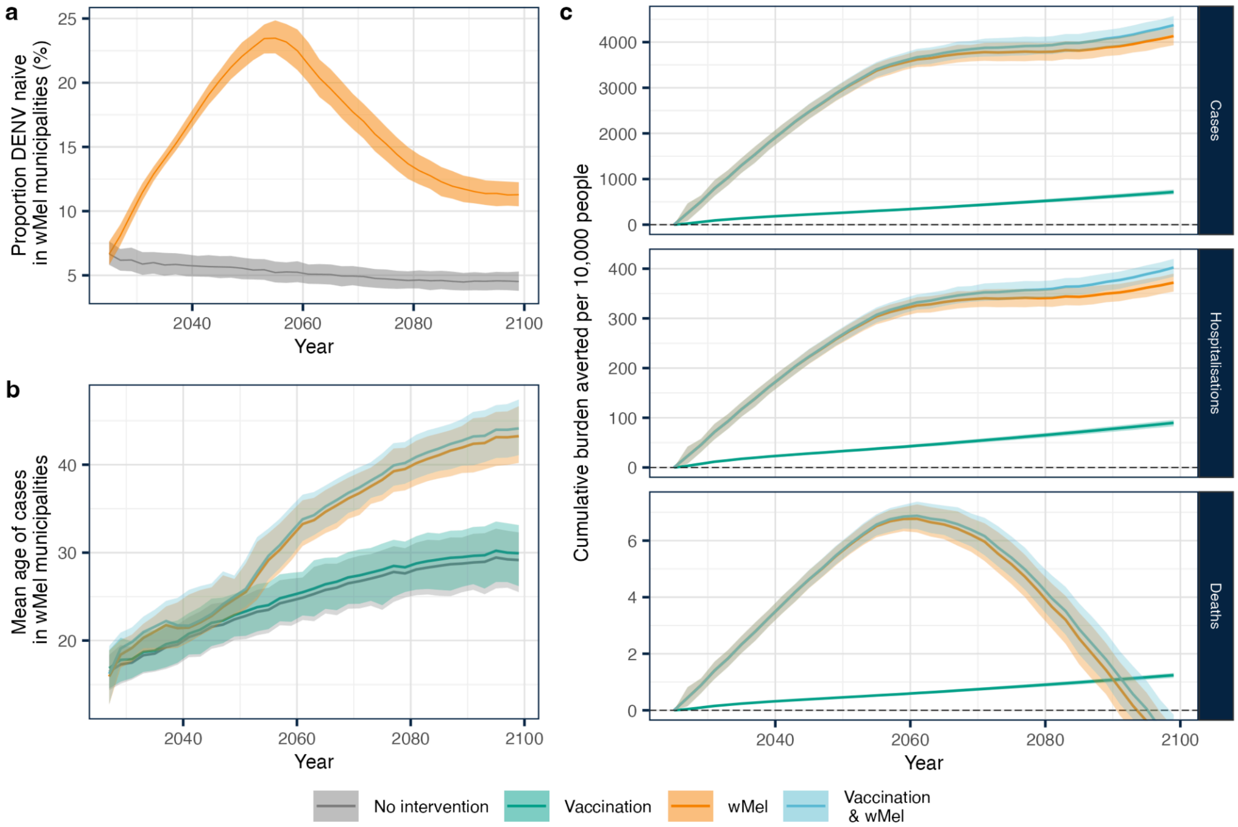


Supplementary Figure 10: Long-term impacts of *w*Mel releases on immunity and public health burden in release areas until 2100

Panel a shows the proportion of individuals living in areas targeted for *w*Mel releases who are DENV naive (seronegative) in a no intervention and *w*Mel scenario. Note that, as we model vaccination as protecting against symptomatic disease and not infection, the proportion DENV-naive in the Vaccination scenario and Vaccination & wMel scenario would be identical to the No intervention scenario and *w*Mel scenario, respectively. Panel b shows the mean age of cases in areas targeted for *w*Mel releases under each scenario. Panel d shows the cumulative cases, hospitalisation and deaths averted per 10,000 population in *w*Mel release areas.
